# A statistical framework for evaluating the repeatability and reproducibility of large language models

**DOI:** 10.1101/2025.08.06.25333170

**Authors:** Cathy Shyr, Boyu Ren, Chih-Yuan Hsu, Chao Yan, Rory J. Tinker, Thomas A. Cassini, Rizwan Hamid, Adam Wright, Lisa Bastarache, Josh F. Peterson, Bradley A. Malin, Hua Xu

## Abstract

**Objective:** Systematic evaluation of variability in large language model (LLM)-generated outputs is critical for assessing their reliability. We developed a regulatory-informed statistical framework to quantify LLM repeatability and reproducibility.

**Materials and Methods:** Using definitions from U.S. Food and Drug Administration draft guidance for AI-enabled medical software, our framework quantifies LLM repeatability and reproducibility along two complementary dimensions: semantic and internal. Semantic metrics quantify variability in the meaning of LLM-generated outputs, and internal metrics quantify variability in token-level probability distributions during text generation. We applied the framework to diagnostic reasoning questions across multiple LLMs and prompting strategies using two datasets: 518 U.S. Medical Licensing Examination (USMLE) questions and 90 real-world rare disease cases from the Undiagnosed Diseases Network (UDN).

**Results:** LLM repeatability and reproducibility varied by dataset, prompting strategy, and model. Prompts eliciting Bayesian diagnostic reasoning achieved significantly higher semantic repeatability than other prompting strategies (***p* < 0.001**). LLM repeatability and reproducibility were generally not associated with diagnostic accuracy.

**Discussion:** LLM repeatability and reproducibility depend not only on the model, but also on the prompting strategy used to elicit reasoning. A model may produce a correct response in a single run but not consistently reproduce that response across repeated runs.

**Conclusion:** We developed a general framework for quantifying LLM repeatability and reproducibility across models, prompting strategies, and biomedical tasks. The metrics in this framework can be used to systematically compare output variability across different LLMs, prompts, model configurations, or evaluation datasets.

## 1 INTRODUCTION

Large language models (LLMs) are increasingly applied in biomedical settings and have demonstrated promising performance across clinical applications, including generating clinical documentation, responding to patient messages, automating phenotyping, and supporting clinical decision-making [1–13]. Evaluation of LLMs has largely focused on task-level metrics such as accuracy. However, accuracy alone does not capture how consistently a model behaves across repeated runs. Because LLMs generate text by sampling tokens from probability distributions, identical prompts can yield different outputs across runs [14]. Therefore, a model may produce a correct response in a single run but fail to reproduce that response consistently across repeated runs, potentially complicating interpretation of model outputs in clinical settings [15]. Despite the distinction between accuracy and output variability, systematic approaches for quantifying variability of LLM outputs remain limited, leaving an important gap in the comprehensive evaluation of model performance.

The importance of measuring variability of LLM outputs has been increasingly emphasized by regulatory agencies and research communities. Draft guidance from the U.S. Food and Drug Administration (FDA) on AI-enabled medical software recommends quantifying model variability based on repeatability and reproducibility [16]. Medical journals recommend researchers to assess variability as a critical component when evaluating the application of LLMs in clinical research [17]. In addition, reporting standards, such as CONSORT-AI and DECIDE-AI, emphasize reproducibility and rigorous evaluation of AI systems in clinical studies [18, 19]. Collectively, these recommendations underscore the importance of systematically assessing variability as part of the comprehensive evaluation of LLM performance.

Variability is common in biomedical settings, as different clinicians may document cases differently or arrive at different diagnoses depending on their training or specialty [20]. The key distinction is that clinicians can explain their reasoning and contextualize such differences, whereas LLMs lack transparent indicators of how much their responses vary across repeated runs [21]. For example, an LLM used for diagnostic support may correctly identify a patient’s diagnosis in one run but generate a different set of candidate diagnoses when given the same patient case again. Both outputs may appear plausible, yet their inconsistency can complicate interpretation and reduce confidence in the model’s recommendations. Without systematic methods to quantify this variability, it is difficult for users to assess the consistency of model outputs across repeated use. Quantifying variability is therefore critical for evaluating the robustness of LLMs and complements accuracy-based metrics to provide a more comprehensive assessment of model performance.

Existing metrics commonly used for evaluating LLM-generated outputs, including BLEU, ROUGE, and BERTScore, were developed for natural language generation tasks such as machine translation and summarization [22–24]. These metrics assess similarity between a model’s output and a reference text based on lexical overlap or semantic similarity. While such metrics are useful for evaluating the accuracy or relevance of model-generated text relative to a ground truth reference, they were not designed to quantify variability across repeated runs. In parallel, prompt engineering has received increasing attention, with studies proposing different strategies such as chain-of-thought prompting for complex reasoning, few-shot prompting for task adaptation, and self-consistency prompting for improving reasoning accuracy [25–29]. However, these approaches typically evaluate prompts based on accuracy-related performance metrics rather than run-to-run variability. As a result, existing methods provide limited insight into how consistently an LLM produces outputs across multiple runs, which is critical for assessing model robustness and reliability in biomedical applications.

To address this gap, our objective was to develop a regulatory-informed statistical framework for quantifying variability of LLM outputs. Consistent with FDA recommendations for evaluating AI-enabled software, our framework operationalizes the evaluation of repeatability, defined as the agreement of model outputs under identical conditions, and reproducibility, agreement under pre-specified, different conditions (e.g., different users or experimental setup) [16]. Within these concepts, our framework further defines two complementary dimensions of variability, semantic and internal. These dimensions yield a total of four metrics: 1) Semantic Repeatability, 2) Internal Repeatability, 3) Semantic Reproducibility, and 4) Internal Reproducibility. Semantic metrics quantify variability in the meaning of LLM outputs across runs, whereas internal metrics quantify variability in the model’s token-level probability distributions, which reflects the stability of the model’s text-generation process. Importantly, our framework is agnostic to both the prompt and the specific model within the class of autoregressive LLMs, which include most modern systems such as ChatGPT, Claude, LLaMA, Qwen, and Gemini [30–32]. We demonstrate the framework using diagnostic reasoning as a use case, evaluating LLM outputs on U.S. Medical Licensing Examination questions and real-world rare disease patient cases from the Undiagnosed Diseases Network. More broadly, the proposed framework provides a systematic approach for quantifying variability of LLM outputs, enabling more comprehensive evaluation of model performance in biomedical research and applications.

## 2 MATERIALS AND METHODS

### 2.1 Definitions of Repeatability and Reproducibility

Motivated by the U.S. FDA’s draft guidance on AI-enabled medical software, which recommends evaluation of both repeatability and reproducibility in AI systems, our framework defines and operationalizes four metrics: 1) Semantic Repeatability, 2) Internal Repeatability, 3) Semantic Reproducibility, and 4) Internal Reproducibility (**Figure 1**). Repeatability is defined by the FDA as “the closeness of agreement of repeated measurements taken under the same conditions.” [16] In our framework, repeatability refers to generating LLM outputs across repeated runs using the same model, prompt, and generation parameters, and measuring the agreement across these outputs. Reproducibility, in contrast, is defined by the FDA as “the closeness of agreement of repeated measurements taken under different, pre-specified conditions.” [16] In our framework, reproducibility refers to generating outputs under controlled variation in one or more experimental conditions while holding other factors constant. These conditions may include, for example, different prompts, users, or care settings.

**Fig. 1:**
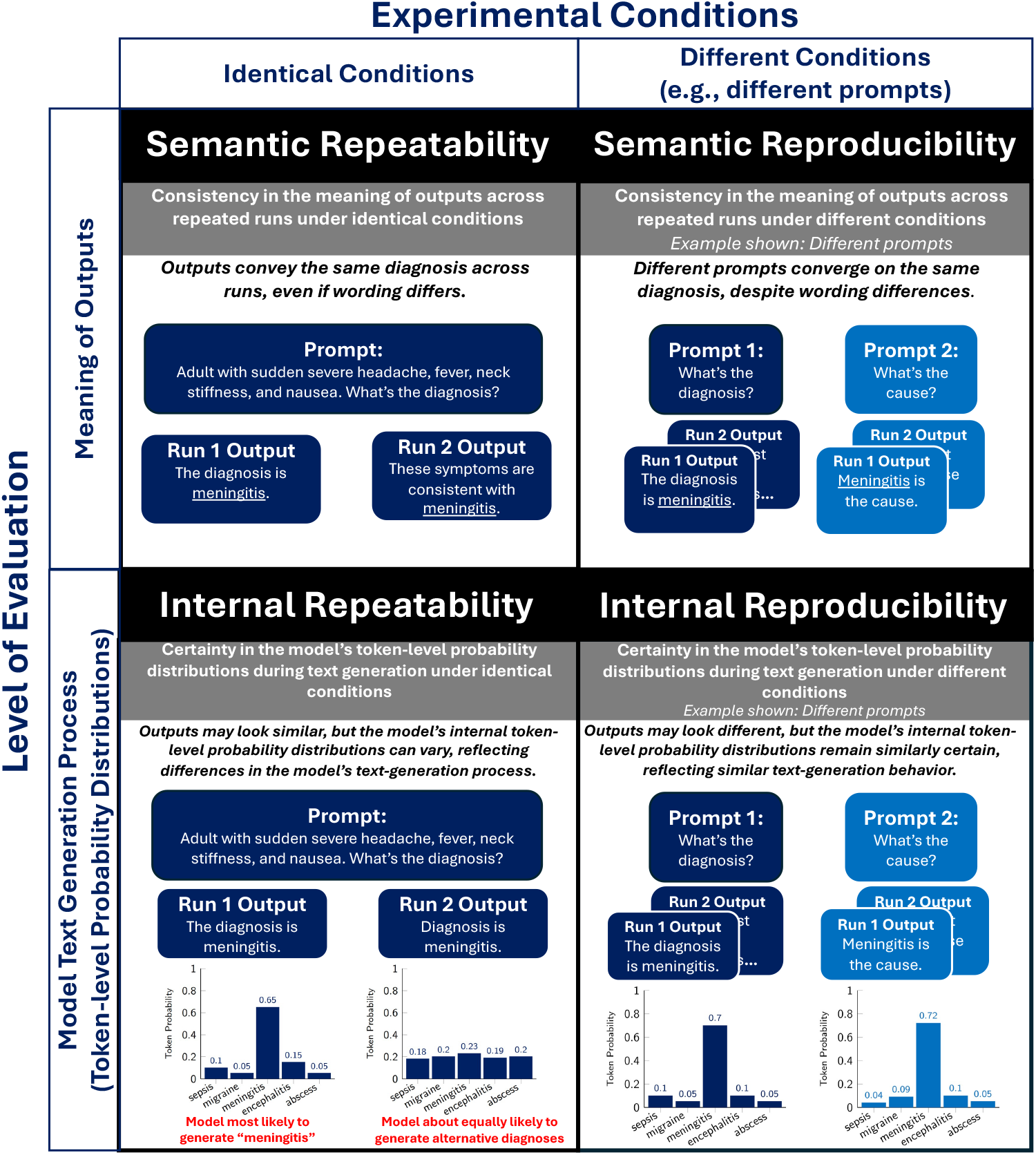
Overview of LLM variability metrics. (1) Semantic Repeatability: consistency of meaning across repeated runs under identical conditions. (2) Semantic Reproducibility: consistency of meaning across repeated runs under different, prespecified conditions (illustrated here using different prompts as an example). (3) Internal Repeatability: certainty in the token-level probability distributions during text generation under identical conditions. (4) Internal Reproducibility: certainty in the token-level probability distributions during text generation under different, prespecified conditions (illustrated here using different prompts as an example).

Within both repeatability and reproducibility, we evaluate LLM outputs along two complementary dimensions: semantic and internal metrics (**Figure 1**). Semantic metrics quantify variability in the meaning of outputs across repeated runs, capturing a clinically relevant property that assesses meaning rather than surface-level wording differences. Internal metrics, by contrast, quantify variability in the token-level probability distributions that underlie LLM text generation. During text generation, autoregressive LLMs (e.g., ChatGPT, LLaMA, etc.) sample the next token from a probability distribution conditioned on prior context. Consequently, two outputs may appear nearly identical on the surface (e.g., “The diagnosis is meningitis” and “Diagnosis is meningitis”) yet differ in their underlying token-level probability distributions during text generation. For example, one run may assign a high probability to the diagnosis “meningitis” while another assigns roughly equal probabilities across several candidate diagnoses (**Figure 1**). Such differences reflect variability in the LLM’s internal text-generation process.

### 2.2 Statistical Framework for Evaluating LLM Repeatability and Reproducibility

In this section, we formally define the four proposed metrics of LLM variability. Let *X* denote the input prompt and 𝒱 the LLM’s output vocabulary (i.e., the set of all possible output tokens). Let *Y*_*r,i*_ ∈ 𝒱 denote the output token generated by the LLM at position *i* = 1 …, *L*_*r*_, where *L*_*r*_ is the length of the output sequence in run *r* = 1 …, *R*, and *R* is the total number of runs. We denote the full output sequence generated by an LLM in run *r* as 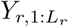.

#### 2.2.1 Semantic Repeatability Score

The Semantic Repeatability Score quantifies the consistency in the meaning of LLM outputs across repeated runs under identical conditions (e.g., same prompt, LLM, text-generation parameters, etc.). Formally, let 𝒱^∗^ denote the set of all finite-length token sequences over the vocabulary 𝒱. Let ℰ: 𝒱^∗^ *→*ℝ be an embedding function that maps an output sequence to a *d*-dimensional vector representation. The embedding function may be implemented using any suitable text embedding model and is not required to be the same model that generated the outputs. For each run *r* = 1, …, *R*, we compute the embedding vector 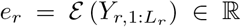. We define the Semantic Repeatability Score in **Definition 1**.

##### Definition 1.

**Semantic Repeatability Score**. The Semantic Repeatability Score quantifies the consistency in the meaning of LLM outputs across repeated runs under identical conditions. Formally, it’s defined as the average pairwise cosine similarity between the embedding representations of LLM outputs across runs:

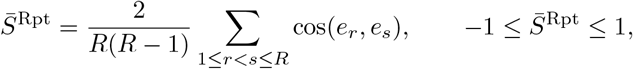

For interpretability, we rescale 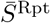 to the interval [0, 1]:

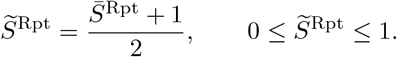

Larger values of 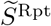 indicate greater semantic consistency across LLM outputs (i.e., larger = more repeatable).

#### 2.2.2 Internal Repeatability Score

The Internal Repeatability Score quantifies the certainty of an autoregressive LLM’s token-level probability distributions during text generation under identical conditions (e.g., same prompt, LLM, text-generation parameters, etc.). Unlike the Semantic Repeatability Score, which evaluates the semantic similarity of final outputs, the Internal Repeatability Score characterizes the internal probabilistic sampling mechanisms used by autoregressive LLMs during text generation.

To define the Internal Repeatability Score, we first outline the text-generation process of autoregressive LLMs. Autoregressive LLMs, which include most modern systems such as ChatGPT, Gemini, Qwen, and LLaMA, generate text sequentially by sampling tokens one at a time [30–32]. At each step, the model produces a probability distribution over its vocabulary and samples the next token from that distribution, conditioned on the prompt and previously generated tokens. Formally, this process can be described as follows:

**Step 1. (User provides a prompt):** The user provides an input prompt *X*.

**Step 2. (Logit computation):** At position *i* of the generated output sequence in run *r*, the LLM produces a vector of logits over the vocabulary 𝒱

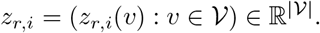

These logits are produced by the LLM decoding function

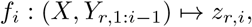

where *Y*_*r*,1:*i*−1_ denotes the history of tokens generated up to position *i −*1. At the first position (*i* = 1), the logits depend only on the user’s prompt:

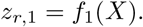

At subsequent positions (*i* = 2, …, *L*_*r*_), the logits depend on both the prompt and previously generated tokens:

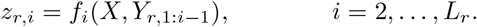

**Step 3. (Conversion to token-level probability distributions):** Let *T* denote the user-specified temperature parameter in LLMs that controls the randomness of token sampling. The logits are transformed into a probability distribution over the vocabulary using the temperature-scaled softmax function:

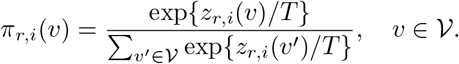

Larger values of *T* produce flatter probability distributions over all possible tokens (i.e., more uncertainty), whereas smaller values produce more peaked distributions (i.e., specific tokens have a higher probability of being sampled).

**Step 4. (Top-***k* **truncation):** To focus on the tokens most likely to be sampled and reduce the influence of extremely low-probability tokens, users can specify the parameter, top-*k*, to truncate the probability distribution *π*_*r,i*_ (·) to the *k* tokens with the highest probabilities of being sampled. Let *K*_*r,i*_ ⊂𝒱 denote this set of tokens. The top-*k* truncated probability distribution is

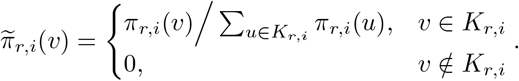

**Step 5. (Token sampling):** The next output token is sampled from the truncated probability distribution 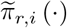.

Steps 2 through 5 repeats until the LLM generates the last token in the output sequence. To quantify the uncertainty of these token-level probability distributions, we calculate the Shannon entropy of the truncated distribution at each position:

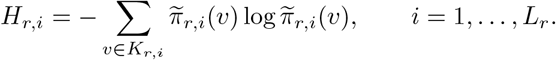

Lower entropy corresponds to more peaked (more certain) token-level probability distributions, whereas higher entropy indicates greater uncertainty across candidate tokens. We average entropy across output positions to obtain the run-level entropy,

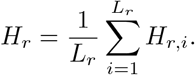

We define the Internal Repeatability Score in **Definition 2**

##### Definition 2.

**Internal Repeatability Score**. The Internal Repeatability Score quantifies the certainty of an LLM’s token-level probability distributions during text generation under identical conditions (e.g., same prompt, model, and generation parameters). To define it, we first compute the mean entropy across *R* independent runs as

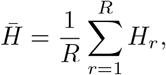

where *H*_*r*_ is the run-level entropy in run *r*. Because the entropy of a distribution supported on at most *k* tokens is bounded above by log_2_ *k*, the mean entropy satisfies 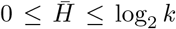. We then rescale 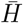 to the interval [0, 1] to obtain the Internal Repeatability Score,

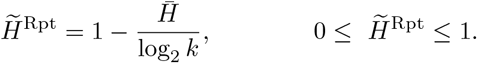

Larger values of 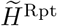 indicate greater certainty in the model’s token-level probability distributions during text generation under identical conditions (i.e., larger = more repeatable).

#### 2.2.3 Semantic Reproducibility Score

The Semantic Reproducibility Score quantifies the consistency in the meaning of LLM outputs across repeated runs under different, pre-specified experimental conditions. These conditions may include, for example, different prompts, users, LLMs, or care settings. Let *P* denote the number of experimental conditions considered (e.g., *P* different prompts). For each condition *p* = 1, …, *P*, we generate *R* independent runs and compute the embedding of each generated output using the embedding function ℰ. Formally, for run *r* under condition *p*, we compute the embedding

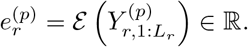

We then compute the average embedding across runs for each condition *p*,

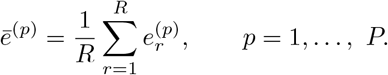

Using these condition-level embeddings, we define the Semantic Reproducibility Score in **Definition 3**.

##### Definition 3.

**Semantic Reproducibility Score**. The Semantic Reproducibility Score quantifies the consistency in the meaning of LLM outputs across repeated runs under different, pre-specified experimental conditions. Formally, it’s defined as the average pairwise cosine similarity between the mean embedding representations of outputs across experimental conditions:

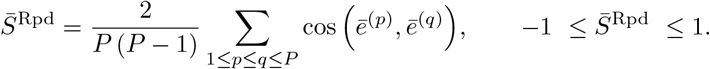

For interpretability, we rescale 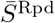 to the interval [0, 1]:

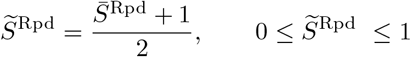

Larger values of 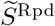 indicate greater consistency in the semantic meaning of LLM outputs across experimental conditions (i.e., larger = more reproducible).

#### 2.2.4 Internal Reproducibility Score

The Internal Reproducibility Score quantifies the consistency in the certainty of an autoregressive LLM’s token-level probability distributions during text generation under different, pre-specified experimental conditions. Let *P* denote the number of experimental conditions considered (e.g., *P* different prompts). For each condition *p* = 1, …, *P*, we generate *R* independent runs and compute the run-level entropy 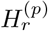 using the same procedure defined for the Internal Repeatability Score. Here, 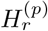 denotes the mean token-level entropy of the truncated probability distributions generated during run *r* under condition *p*. We then compute the average run-level entropy across runs for each condition,

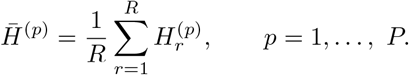

Using these condition-level entropy estimates, we define the Internal Reproducibility Score in **Definition 4**.

##### Definition 4.

**Internal Reproducibility Score**. The Internal Reproducibility Score quantifies the certainty of token-level probability distributions during text generation under different, pre-specified experimental conditions. Formally, it is defined as the average pairwise difference in mean entropy between conditions, rescaled to the interval [0, 1] for interpretability:

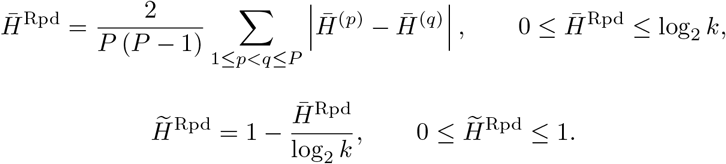

where 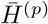 denotes the mean run-level entropy under experimental condition *p*. Larger values of 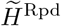 indicate greater consistency in the certainty of the model’s token-level probability distributions across experimental conditions (i.e., larger = more reproducible).

### 2.3 Empirical Evaluation of LLM Diagnostic Reasoning

To demonstrate the practical application of the proposed repeatability and reproducibility metrics, we performed an empirical evaluation of LLM diagnostic reasoning across multiple clinical datasets, prompting strategies, and model architectures. In this empirical use case, reproducibility was operationalized using different diagnostic reasoning prompts. The proposed framework, however, is general and can be applied to quantify reproducibility across any pre-specified experimental condition, such as different LLMs, users, or care settings.

#### 2.3.1 Diagnostic Reasoning Prompts

We evaluated LLM repeatability and reproducibility across five chain-of-thought (CoT) diagnostic reasoning prompts developed and validated by Savage *et al* [33]. These prompts were designed to elicit distinct diagnostic reasoning strategies used in clinical practice, including intuitive reasoning, analytic reasoning, and probabilistic reasoning (**Table 1**). The full prompts are provided in the **Supplementary Materials**.

**Table 1:**
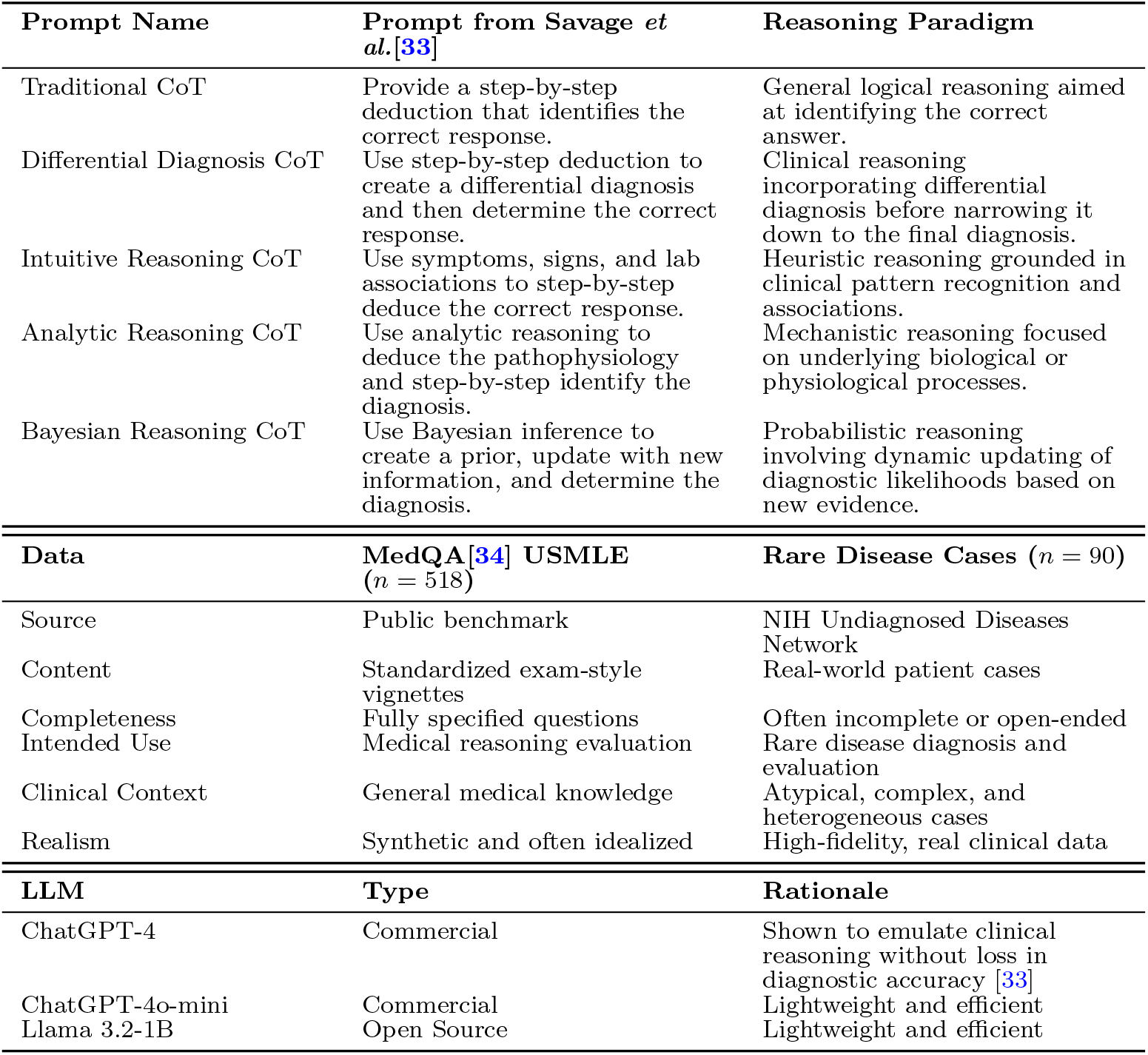
Overview of empirical evaluation. CoT = chain of thought; USMLE = U.S. Medical Licensing Examination; NIH = National Institutes of Health; LLM = Large language model

#### 2.3.2 Datasets

To evaluate repeatability and reproducibility across diverse clinical contexts, we selected two complementary datasets: 1) a standardized benchmark for medical knowledge (MedQA), and 2) real-world rare disease cases from the Undiagnosed Diseases Network (UDN) [34, 35].

MedQA is a publicly available benchmark consisting of diagnostic clinical vignettes derived from U.S. Medical Licensing Examination (USMLE) questions [34]. Following Savage *et al*., we used the same set of 518 questions, reformulated from multiple-choice format into free-response prompts [33]. We focused on Step 2 and Step 3 cases, which emphasize clinical reasoning over rote recall. These vignettes are fully specified and standardized, making them well-suited for controlled benchmarking of diagnostic reasoning. However, they may be idealized relative to real-world clinical scenarios. An example is shown in **Table 2**. All questions are publicly available and provided in **Supplementary Data 1** of Savage *et al* [33].

**Table 2:**
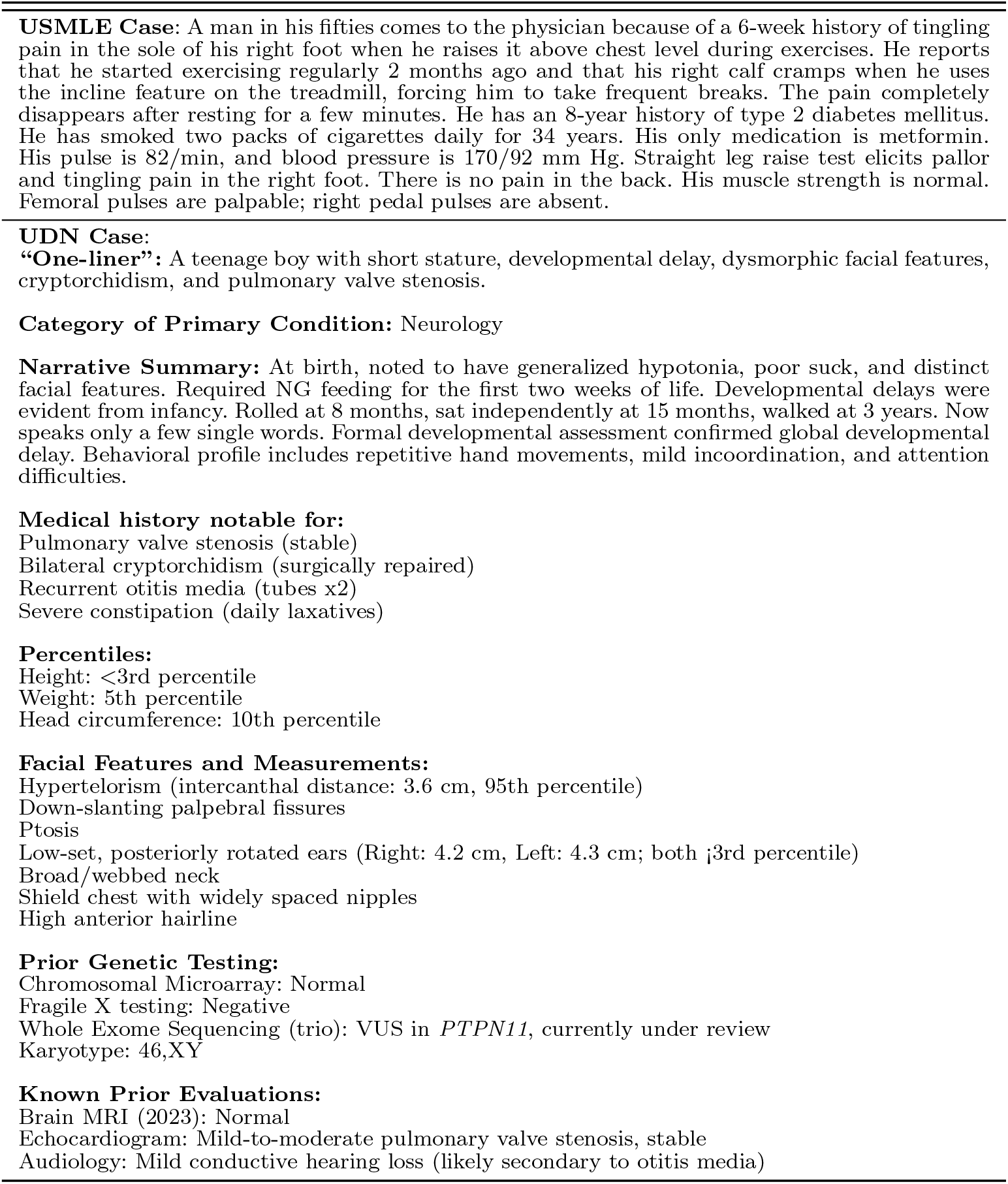
Example of USMLE and UDN clinical cases. USMLE = U.S. Medical Licensing Examination; UDN = Undiagnosed Diseases Network.

To complement MedQA and address concerns that public benchmarks may have appeared during LLM pre-training, we additionally analyzed 90 non-public rare disease cases from the Vanderbilt University Medical Center (VUMC) site of the UDN [36]. In contrast to the exam-style vignettes of USMLE, UDN patients often present with complex and heterogeneous phenotypes, frequently involving multi-system manifestations, non-diagnostic clinical test results, and atypical genotypes that are challenging to interpret (e.g., variants of uncertain significance) [37, 38]. They also may have symptoms or test results included in the case data that are unrelated to their final diagnosis. Each UDN case is summarized as a multi-paragraph narrative detailing the patient’s medical history and prior workup. An illustrative example is shown in **Table 2**. This study was approved by the VUMC Institutional Review Board (IRB# 172005).

#### 2.3.3 LLMs

We evaluated three LLMs selected to represent a range of model sizes (**Table 1**): 1) ChatGPT-4 (OpenAI) is a commercial LLM that has previously demonstrated strong diagnostic reasoning performance on the same USMLE dataset used in this study [33]; 2) ChatGPT-4o-mini (OpenAI) is a smaller, more cost-efficient variant of GPT-4 designed for more lightweight deployments. Including this model allows us to evaluate repeatability and reproducibility under a lightweight, resource-efficient setting; 3) LLaMA 3.2-1B (Meta AI) is an open-source, lightweight model that provides a computationally efficient alternative to commercial systems [30].

#### 2.3.4 Evaluation Setup and Parameter Settings

ChatGPT models were accessed via the Microsoft Azure OpenAI application programming interface, and LLaMA 3.2-1B was deployed on a secure VUMC server. To ensure patient privacy, all evaluations involving UDN cases were conducted using institutionally approved secure computing environments. For all runs, we set the temperature *T* to 0.5 and top-*k* to 30. This parameter configuration was chosen to balance determinism and diversity so that both repeatability and reproducibility could be meaningfully evaluated. Extremely deterministic settings could artificially inflate repeatability, whereas highly stochastic settings could exaggerate variability. Although fixed in our evaluation, the proposed framework is agnostic to parameter choice and can be applied under any parameter configuration appropriate to the user’s application.

#### 2.3.5 Statistical Analysis

We used two-sided multivariate Kruskal-Wallis tests at the 0.05 level to assess differences in repeatability and reproducibility scores across diagnostic reasoning prompts, datasets (USMLE vs. UDN), and LLMs. When the result from a Kruskal-Wallis test was statistically significant, we performed pairwise post-hoc comparisons using Dunn’s test. To account for multiple testing, p-values were adjusted using the Holm-Bonferroni method. Adjusted p-values less than 0.05 were considered statistically significant.

## 3 RESULTS

### 3.1 Results from the Empirical Evaluation of LLM Diagnostic Reasoning

Across the five diagnostic reasoning prompts, we evaluated 518 USMLE cases and 90 UDN cases using three LLMs with *R* = 100 independent runs per promptcase-LLM combination, resulting in 912, 000 total generations. Repeatability metrics (Semantic Repeatability and Internal Repeatability) were computed within each prompt-case-LLM combination across repeated runs. Reproducibility metrics (Semantic Reproducibility and Internal Reproducibility) were computed across prompts for each case-LLM combination.

#### 3.1.1 Repeatability of LLM Diagnostic Reasoning

Overall, semantic repeatability varied across diagnostic reasoning prompts, models, and datasets (**Figure 2**). For ChatGPT-4, the Bayesian CoT prompt tended to produce higher Semantic Repeatability Scores across both USMLE and UDN cases. Differences across prompts were statistically significant for ChatGPT-4o-mini on USMLE cases (*p* < 0.001). In contrast, internal repeatability showed relatively little variation across models and prompting strategies. One exception was ChatGPT-4o-mini, where Traditional CoT and Bayesian CoT prompts had statistically significantly lower Internal Repeatability Scores compared with other prompting strategies (adjusted *p* < 0.001). Across all models, repeatability scores for UDN cases were more tightly clustered across prompting strategies than for USMLE cases (**Figure 2**), indicating less variation in repeatability across prompts. We provide all repeatability scores in **Table S1**.

**Fig. 2:**
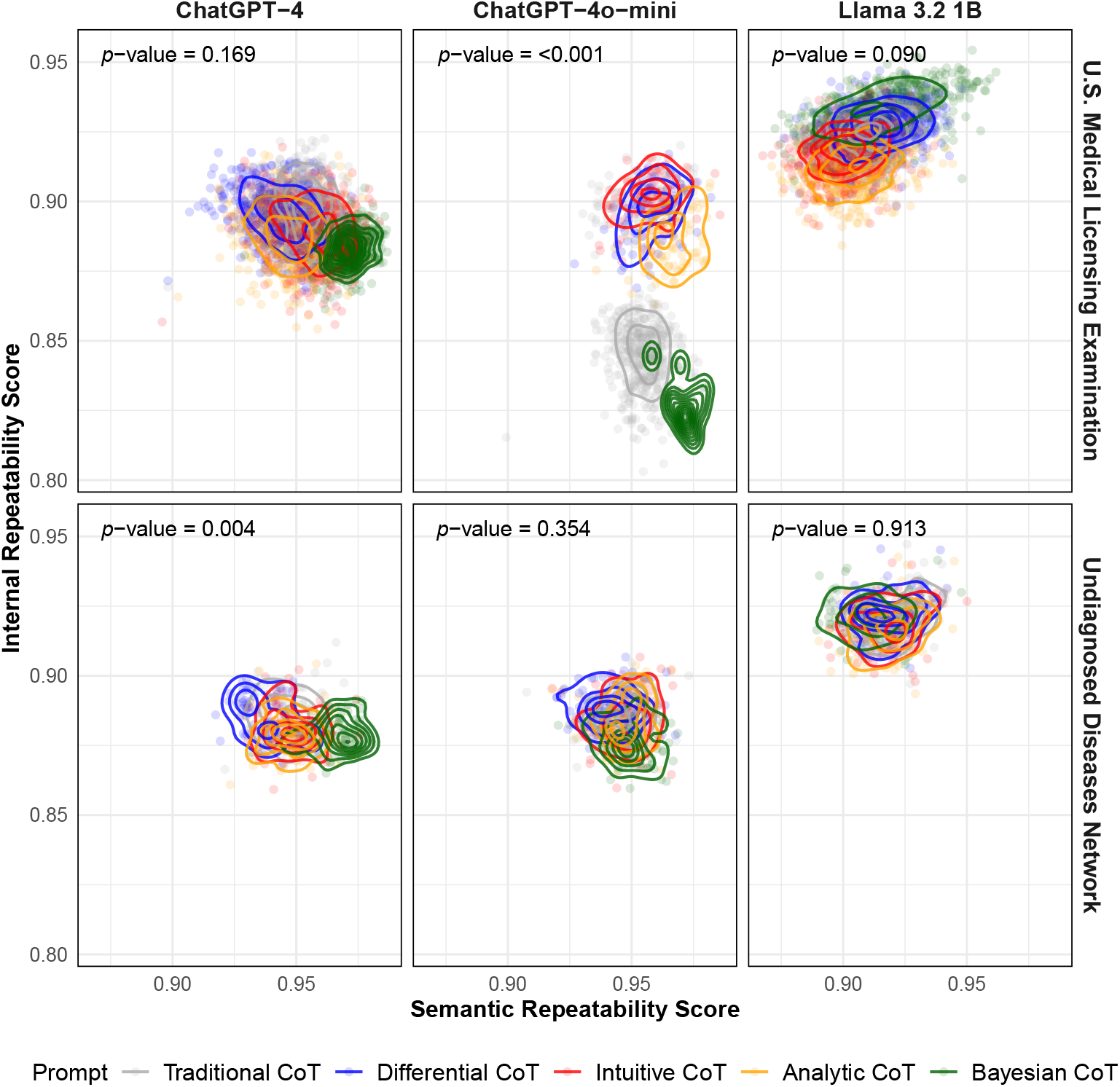
Semantic and Internal Repeatability Scores by large language model, dataset, and prompt. Each dot represents a single clinical case evaluated under a specific prompting strategy, plotted by its Semantic Repeatability Score (x-axis) and Internal Repeatability Score (y-axis). Contour lines represent two-dimensional density estimates for each prompting strategy. Panels are stratified by model (columns) and dataset (rows). *P*-values are calculated based on a two-sided multivariate Kruskal–Wallis test at the 0.05 level. Higher values indicate greater repeatability for both metrics. *CoT = chain-of-thought; UDN = Undiagnosed Diseases Network*.

#### 3.1.2 Reproducibility of LLM Diagnostic Reasoning

In this empirical evaluation, reproducibility was assessed across different diagnostic reasoning prompts (**Table 1**). Across all three LLMs, reproducibility scores varied less for UDN than for USMLE cases (**Figure 3**). On the USMLE cases, ChatGPT-4o-mini achieved higher Internal Reproducibility Scores compared to the other LLMs (adjusted *p* < 0.001). In contrast, LLaMA 3.2-1B exhibited higher Semantic Reproducibility Scores (adjusted *p* < 0.001). For the UDN cases, Internal Reproducibility Scores were relatively uniform across all three LLMs. However, LLaMA 3.2-1B exhibited higher Semantic Reproducibility scores than the other models (adjusted *p* < 0.001). We provide all reproducibility scores in **Table S1**.

**Fig. 3:**
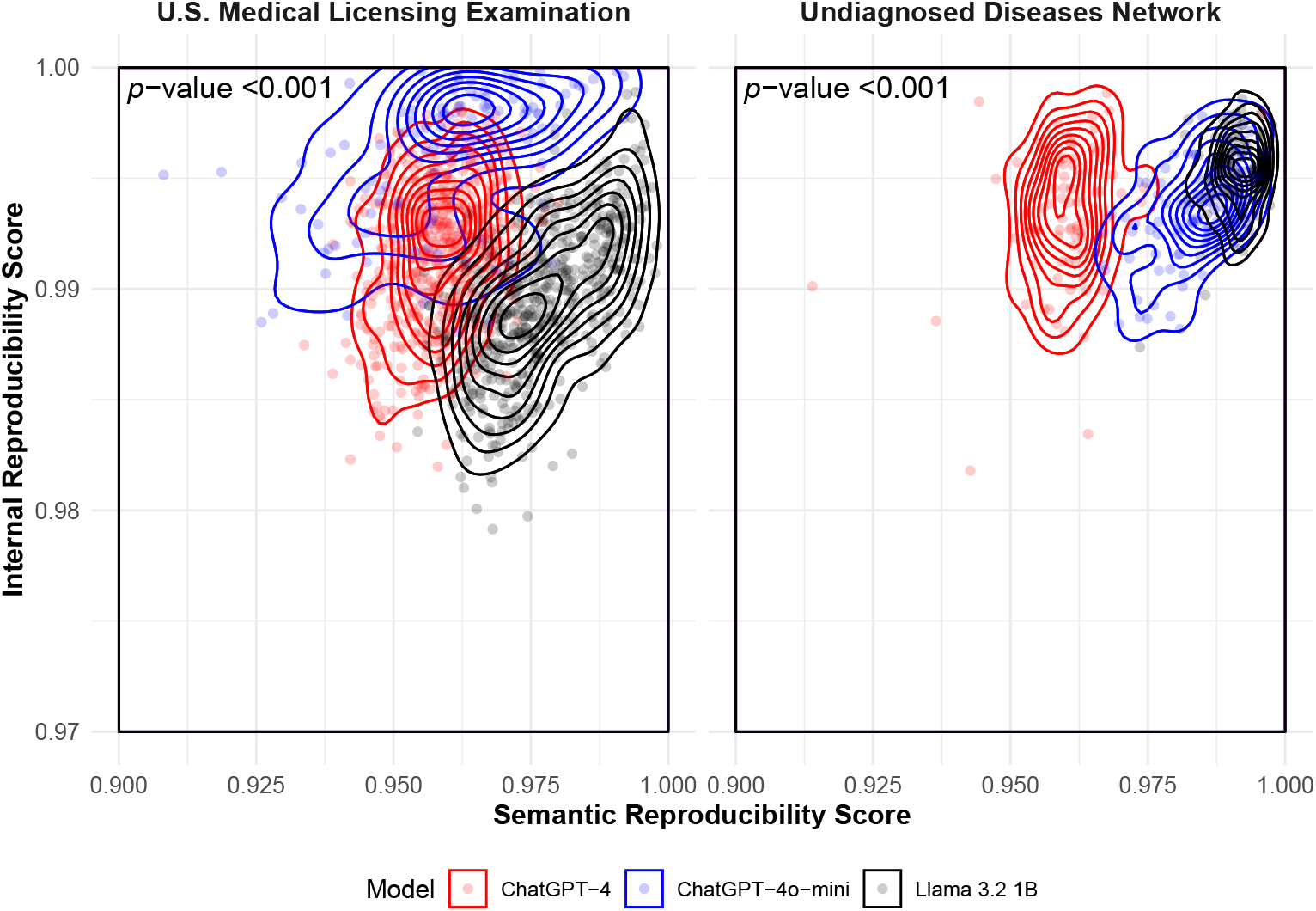
Semantic and Internal Reproducibility Scores by large language model and dataset. Each dot represents a single clinical case evaluated by a model, with Semantic Reproducibility Score (x-axis) and Internal Reproducibility Score (y-axis) computed across the five diagnostic reasoning prompts. Contour lines represent two-dimensional density estimates for each prompting strategy. *P*-values are calculated based on a two-sided multivariate Kruskal–Wallis test at the 0.05 level. Higher values indicate greater reproducibility for both metrics.

#### 3.1.3 Relationship Among LLM Diagnostic Accuracy, Repeatability, and Reproducibility

We further evaluated whether repeatability and reproducibility were associated with diagnostic accuracy using published ground-truth labels for ChatGPT-4 on USMLE cases from Savage *et al* [33]. In that study, physicians manually reviewed ChatGPT-4’s outputs and assigned diagnostic accuracy labels (correctly diagnosed vs. incorrectly diagnosed) for each case (see **Supplementary Information** in Savage *et al*. [33]). Using the same set of USMLE cases and prompts, we generated *R* = 100 outputs per case and computed repeatability and reproducibility scores. We then compared these scores between correctly and incorrectly diagnosed cases. As shown in **Figure 4**, Semantic Repeatability and Internal Repeatability Scores were similar between correctly and incorrectly diagnosed cases across most prompting strategies. For the Traditional CoT, Differential Diagnosis CoT, Analytic CoT, and Bayesian CoT prompts, no statistically significant differences in repeatability scores were observed between the two groups. However, under the Intuitive CoT prompting strategy, correctly diagnosed cases exhibited significantly higher Internal Repeatability Scores than incorrectly diagnosed cases (*p* < 0.001). **Figure 5** shows the corresponding analysis for reproducibility scores. No statistically significant differences in either semantic reproducibility or internal reproducibility were observed between correctly and incorrectly diagnosed cases (*p* = 0.26).

**Fig. 4:**
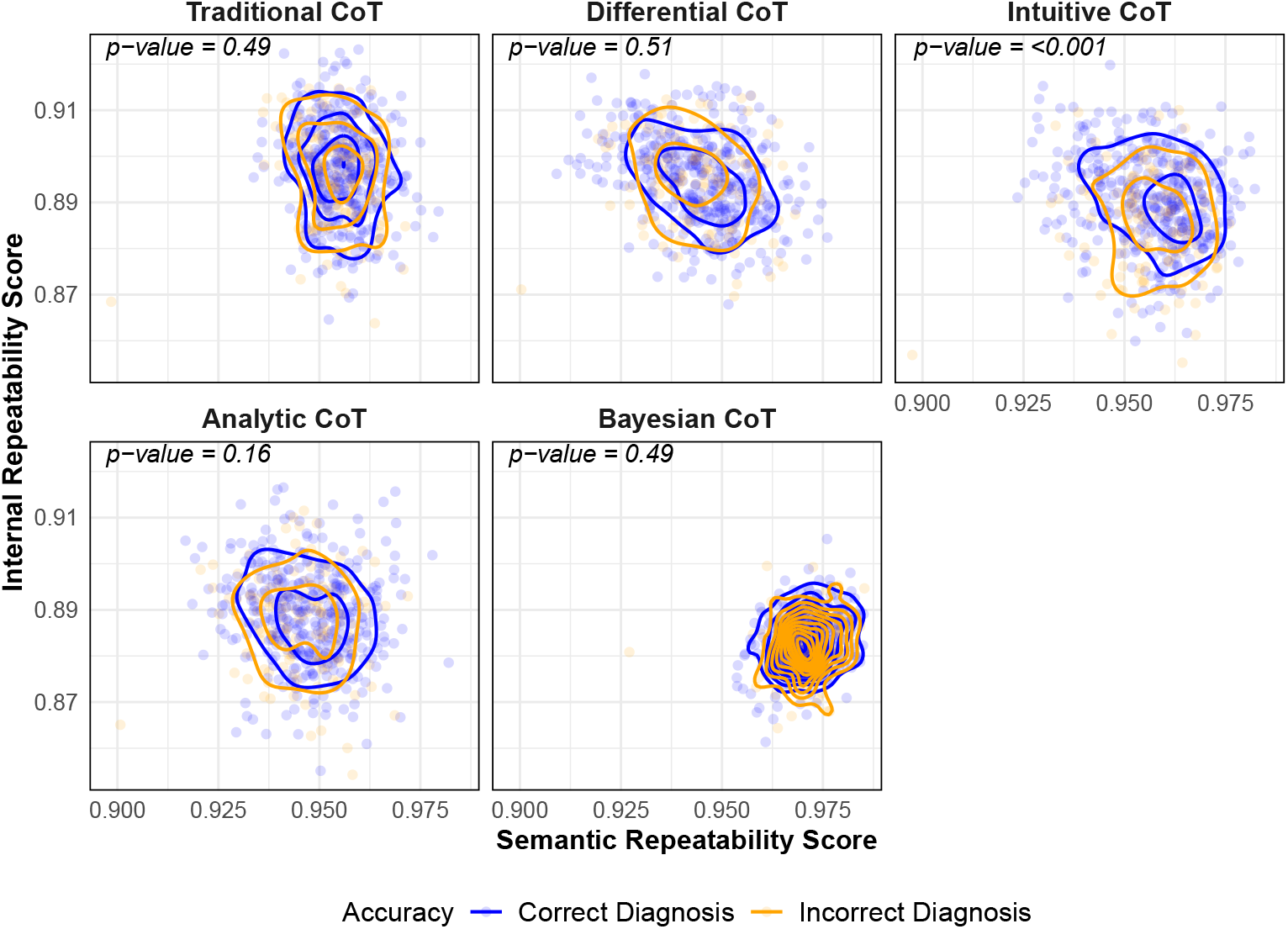
Semantic and Internal Repeatability Scores for ChatGPT-4 on the MedQA (U.S. Medical Licensing Examination) dataset stratified by prompt. Each dot represents a single clinical case evaluated by ChatGPT-4, with Semantic Repeatability Score on the x-axis and Internal Repeatability Score on the y-axis. Contour lines represent two-dimensional density estimates for cases that were correctly diagnosed by ChatGPT-4 (blue) and incorrectly diagnosed by ChatGPT-4 (orange). *P*-values are calculated based on a two-sided multivariate Kruskal-Wallis test at the 0.05 level. Higher values indicate greater repeatability for both metrics.

**Fig. 5:**
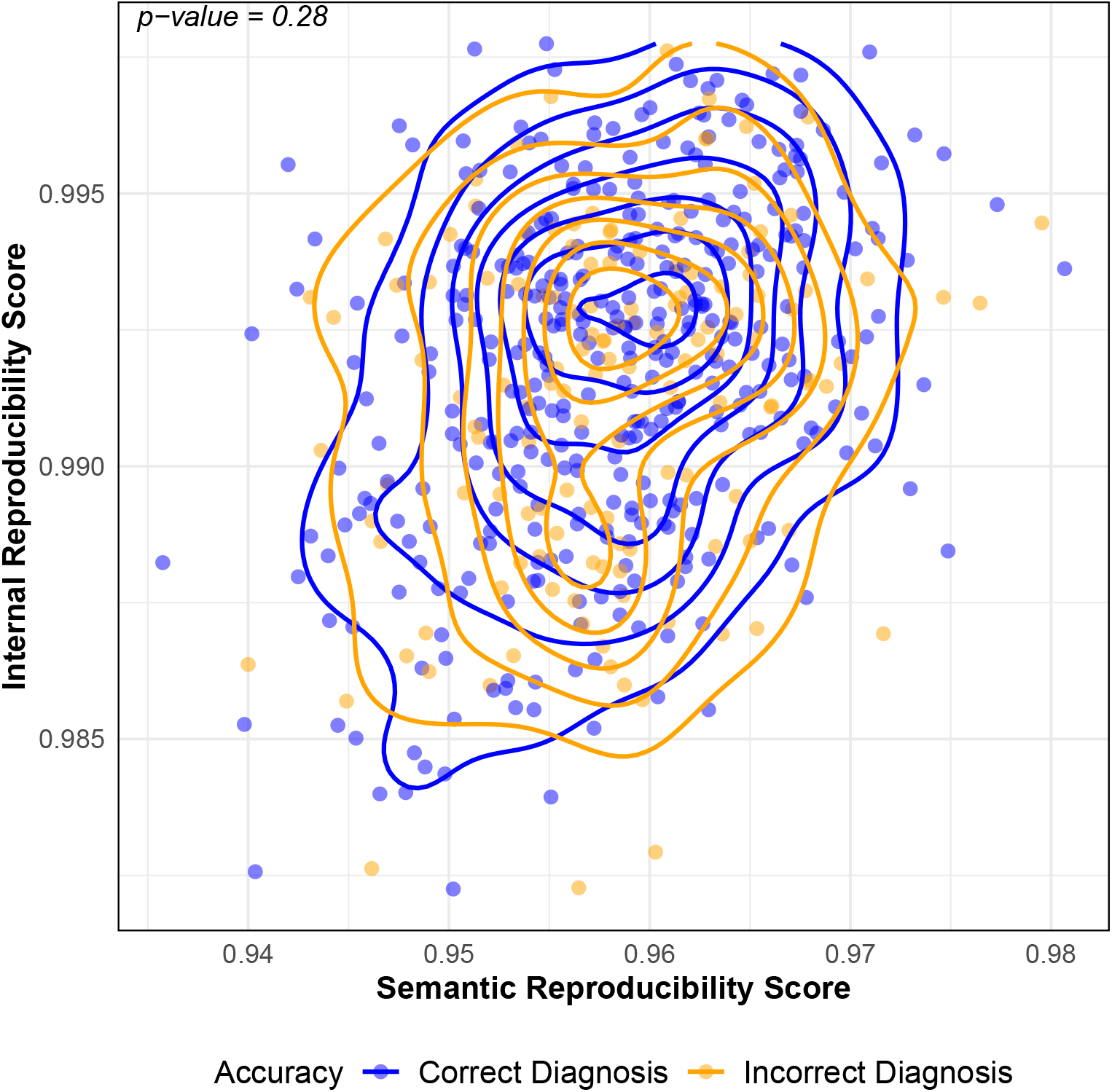
Semantic and Internal Reproducibility Scores for ChatGPT-4 on the MedQA (U.S. Medical Licensing Examination) dataset stratified by prompt. Each dot represents a single clinical case evaluated by ChatGPT-4, with Semantic Reproducibility Score on the x-axis and Internal Reproducibility Score on the y-axis. Contour lines represent two-dimensional density estimates for cases that were correctly diagnosed by ChatGPT-4 (blue) and incorrectly diagnosed by ChatGPT-4 (orange). *P*-values are calculated based on a two-sided multivariate Kruskal-Wallis test at the 0.05 level. Higher values indicate greater reproducibility for both metrics.

## 4 DISCUSSION

We developed a regulatory-informed statistical framework for quantifying the repeatability and reproducibility of LLM outputs. Because the framework is agnostic to the specific LLM, prompting strategy, and evaluation task, it can be applied across diverse biomedical use cases. In practice, researchers can use the repeatability and reproducibility metrics to systematically compare output variability across different LLMs, prompts, model configurations, or evaluation datasets. Rather than defining universal thresholds, these metrics are primarily intended to support relative comparisons. While application-specific thresholds for these metrics may be defined in specific use cases, establishing such thresholds is beyond the scope of this study.

From our empirical evaluation, three key findings emerged. First, repeatability and reproducibility scores varied less for UDN cases than USMLE questions. One possible explanation is that the longer and more detailed narrative structure of real-world patient cases may constrain the range of plausible model responses, resulting in less variability across prompts. Additional studies across other clinical datasets are necessary to determine whether this finding generalizes beyond the specific datasets examined here. Second, prompts invoking Bayesian diagnostic reasoning yielded higher Semantic Repeatability Scores for ChatGPT-4. This finding suggests that repeatability may depend not only on the model itself, but also on the prompting strategy used to elicit reasoning. These findings indicate that repeatability and reproducibility are not fixed properties of an LLM, but depend on the interplay among the model, prompting strategy, and evaluation dataset. Therefore, evaluations based on a single prompting strategy or benchmark dataset may not fully characterize model performance across different settings. Third, repeatability and reproducibility were generally not associated with diagnostic accuracy in our empirical analysis. This observation highlights that correctness and consistency are distinct aspects of model performance. A model may produce a correct output in a single run but fail to reproduce that answer consistently across repeated runs, or conversely produce consistent outputs that are incorrect. Therefore, evaluating repeatability and reproducibility alongside accuracy may provide a more comprehensive characterization of LLM performance.

This work is complementary to prior efforts in natural language processing that quantify and improve LLMs’ output consistency. Raj *et al*. proposed semantic consistency metrics and demonstrated that consistency and accuracy are independent properties, an observation that aligns with our findings [39]. Similarly, Cui *et al*. introduced a divide-and-conquer approach to improve sentence-level consistency, demonstrating that consistency metrics can serve not only as evaluation tools but also as mechanisms for improving model behvaior [40]. Wang *et al*. proposed MONITOR, a framework for assessing factual reliability under prompt variability, further underscoring the limitations of accuracy as the standalone evaluation metric [41]. Complementing these general-domain studies, our work operationalizes repeatability and reproducibility using definitions from FDA recommendations on AI-enabled devices [16], providing a step toward regulatory-informed evaluation of LLMs.

This work has several limitations. First, due to the substantial computational cost of generating repeated outputs across prompts, models, and clinical cases, our empirical evaluation did not exhaustively cover all possible configurations. While we included diverse models, prompts, and datasets to approximate real-world clinical variation, additional contexts should be explored in future work. Second, the Internal Repeatability and Reproducibility Scores rely on access to token-level probabilities. While these probabilities are available in autoregressive LLMs (e.g., ChatGPT, LLaMA), they may not be directly accessible for other LLM architectures. Therefore, the internal metrics proposed in this study are primarily applicable to autoregressive LLMs, whereas the semantic metrics are broadly applicable across LLM architectures. Future work includes how variability metrics can be integrated with human-centered evaluation approaches, such as clinician review or usability assessments, to better understand how variability in model outputs influences clinical interpretation and decision-making [42].

Beyond evaluating model performance, quantifying variability is also important when LLM outputs are treated as study endpoints in biomedical research. Incorporating repeatability and reproducibility into evaluation frameworks may support more rigorous experimental design, model comparison, and interpretation of LLM-generated outputs. Practical considerations, including computational cost, scalability, and integration into clinical workflows, will influence how such evaluation frameworks are applied in practice [4, 42, 43].

## Acknowledgments

The authors are grateful to the patients for participating in the Undiagnosed Diseases Network.

## Data availability

The MedQA (U.S. Medical Licensing Examination) cases used in this study are publicly available in Savage *et al*. [33]’s [Supplementary Data 1](https://doi.org/10.1038/s41746-024-01010-1). The Undiagnosed Diseases Network data used in this study contain sensitive patient information. De-identified patient data, including phenotypic and genomic data, are deposited in the [database of Genotypes and Phenotypes (dbGaP)](https://www.ncbi.nlm.nih.gov/gap/) maintained by the National Institutes of Health. To explore data available in the latest release, visit the [UDN study page in dbGaP](https://www.ncbi.nlm.nih.gov/projects/gap/cgi-bin/study.cgi?study_id=phs001232.v7.p3). Individuals interested in accessing UDN data through dbGaP should submit a data access request. Detailed instructions for this process can be found on the NIH Scientific Data Sharing website: [How to Request and Access Datasets from dbGaP](https://sharing.nih.gov/accessing-data/accessing-genomic-data/how-to-request-and-access-datasets-from-dbgap).

## Code availability

Code used in this study are publicly available at https://github.com/cathyshyr/repeatability and reproducibility of LLMs.

## Author Contributions

Conceptualization: C.S.

Data curation: C.S., B.R., C.H., R.J.T., T.A.C., R.H., L.B.

Formal analysis and interpretation: C.S., B.R., C.H., C.Y., A.W., L.B., J.F.P., B.A.M., H.X.

Methodology: C.S., B.R., C.H. Writing – original draft: C.S.

Writing – review and editing: C.S., B.R., C.H., C.Y., T.J.T., T.A.C., R.H., A.W., L.B., J.F.P., B.A.M., H.X.

Supervision: H.X.

Project administration: C.S., H.X.

Funding acquisition: C.S., R.H.

