## Supplementary Materials for "A statistical framework for evaluating the repeatability and reproducibility of large language models"

**Supplementary Information**

**Supplementary Note 1**

**Full prompts used for MedQA (U.S. Medical Licensing Examination) dataset:** All prompts are from Savage, Thomas, et al. “Diagnostic reasoning prompts reveal the potential for large language model interpretability in medicine.” *NPJ Digital Medicine* 7.1 (2024): 20.

1. Traditional chain-of-thought: “Provide a step-by-step deduction that identifies the correct response.

   Example Question 1:
   Shortly after undergoing a bipolar prosthesis for a displaced femoral neck fracture of the left hip acquired after a fall the day before, an woman in her eighties woman suddenly develops dyspnea. The surgery under general anesthesia with sevoflurane was uneventful, lasting 98 min, during which the patient maintained oxygen saturation readings of 100% on 8 l of oxygen. She has a history of hypertension, osteoporosis, and osteoarthritis of her right knee. Her medications include ramipril, naproxen, ranitidine, and a multivitamin. She appears cyanotic, drowsy, and is oriented only to person. Her temperature is 38.6 °C (101.5 °F), pulse is 135/min, respirations are 36/min, and blood pressure is 155/95 mm Hg. Pulse oximetry on room air shows an oxygen saturation of 81%. There are several scattered petechiae on the anterior chest wall. Laboratory studies show a hemoglobin concentration of 10.5 g/dl, a leukocyte count of 9000/mm3, a platelet count of 145,000/mm3, and a creatine kinase of 190 U/l. An ECG shows sinus tachycardia. What is the most likely diagnosis?

   Example Rationale 1:
   The patient had a surgical repair of a displaced femoral neck fracture. The patient has petechiae. The patient has a new oxygen requirement, meaning they are having difficulty with their breathing. This patient most likely has a fat embolism.

   Example Question 2:
   A man in his fifties man comes to the emergency department because of a dry cough and severe chest pain beginning that morning. Two months ago, he was diagnosed with inferior wall myocardial infarction and was treated with stent implantation of the right coronary artery. He has a history of hypertension and hypercholesterolemia. His medications include aspirin, clopidogrel, atorvastatin, and enalapril. His temperature is 38.5 °C (101.3 °F), pulse is 92/min, respirations are 22/min, and blood pressure is 130/80 mm Hg. Cardiac examination shows a high-pitched scratching sound best heard while sitting upright and during expiration. The remainder of the examination shows no abnormalities. An ECG shows diffuse ST elevations. Serum studies show a troponin I of 0.005 ng/ml (N < 0.01). What is the most likely cause of this patient’s symptoms?

   Example Rationale 2:
   This patient is having chest pain. They recently had a heart attack and has new chest pain, suggesting he may have a problem with his heart. The EKG has diffuse ST elevations and he has a scratching murmur. This patient likely has Dressler Syndrome.

Question:

{Insert MedQA case here}”

2. Differential diagnosis chain-of-thought: “Use step by step deduction to create a differential diagnosis and then use step by step deduction to determine the correct response.

   Example Question 1:
   Shortly after undergoing a bipolar prosthesis for a displaced femoral neck fracture of the left hip acquired after a fall the day before, an woman in her eighties woman suddenly develops dyspnea. The surgery under general anesthesia with sevoflurane was uneventful, lasting 98 min, during which the patient maintained oxygen saturation readings of 100% on 8 l of oxygen. She has a history of hypertension, osteoporosis, and osteoarthritis of her right knee. Her medications include ramipril, naproxen, ranitidine, and a multivitamin. She appears cyanotic, drowsy, and is oriented only to person. Her temperature is 38.6 °C (101.5 °F), pulse is 135/min, respirations are 36/min, and blood pressure is 155/95 mm Hg. Pulse oximetry on room air shows an oxygen saturation of 81%. There are several scattered petechiae on the anterior chest wall. Laboratory studies show a hemoglobin concentration of 10.5 g/dl, a leukocyte count of 9000/mm3, a platelet count of 145,000/mm3, and a creatine kinase of 190 U/l. An ECG shows sinus tachycardia. What is the most likely diagnosis?

   Example Rationale 1:
   This patient has shortness of breath after a long bone surgery. The differential for this patient is pulmonary embolism, fat embolism, myocardial infarction, blood loss, anaphylaxis, or a drug reaction. The patient has petechiae which makes fat embolism more likely. This patient most likely has a fat embolism.

   Example Question 2:
   A man in his fifties man comes to the emergency department because of a dry cough and severe chest pain beginning that morning. Two months ago, he was diagnosed with inferior wall myocardial infarction and was treated with stent implantation of the right coronary artery. He has a history of hypertension and hypercholesterolemia. His medications include aspirin, clopidogrel, atorvastatin, and enalapril. His temperature is 38.5 °C (101.3 °F), pulse is 92/min, respirations are 22/min, and blood pressure is 130/80 mm Hg. Cardiac examination shows a high-pitched scratching sound best heard while sitting upright and during expiration. The remainder of the examination shows no abnormalities. An ECG shows diffuse ST elevations. Serum studies show a troponin I of 0.005 ng/ml (N < 0.01). What is the most likely cause of this patient’s symptoms?

   Example Rationale 2:
   This patient has chest pain with diffuse ST elevations after a recent myocardial infarction. The differential for this patient includes: myocardial infarction, pulmonary embolism, pericarditis, Dressler syndrome, aortic dissection, and costochondritis. This patient likely has a high-pitched scratching sound on auscultation associated with pericarditis and Dressler Syndrome. This patient has diffuse ST elevations associated with Dressler Syndrome. This patient most likely has Dressler Syndrome.

Question:

{Insert MedQA case here}”

3. Intuitive reasoning chain-of-thought: “Use symptoms, signs, and laboratory disease associations to step by step deduce the correct response.

   Example Question 1:
   Shortly after undergoing a bipolar prosthesis for a displaced femoral neck fracture of the left hip acquired after a fall the day before, an woman in her eighties woman suddenly develops dyspnea. The surgery under general anesthesia with sevoflurane was uneventful, lasting 98 min, during which the patient maintained oxygen saturation readings of 100% on 8 l of oxygen. She has a history of hypertension, osteoporosis, and osteoarthritis of her right knee. Her medications include ramipril, naproxen, ranitidine, and a multivitamin. She appears cyanotic, drowsy, and is oriented only to person. Her temperature is 38.6 °C (101.5 °F), pulse is 135/min, respirations are 36/min, and blood pressure is 155/95 mm Hg. Pulse oximetry on room air shows an oxygen saturation of 81%. There are several scattered petechiae on the anterior chest wall. Laboratory studies show a hemoglobin concentration of 10.5 g/dl, a leukocyte count of 9000/mm3, a platelet count of 145,000/mm3, and a creatine kinase of 190 U/l. An ECG shows sinus tachycardia. What is the most likely diagnosis?

   Example Rationale 1:
   This patient has findings of petechiae, altered mental status, shortness of breath, and recent surgery suggesting a diagnosis of fat emboli. The patient most likely has a fat embolism.

   Example Question 2:
   A man in his fifties man comes to the emergency department because of a dry cough and severe chest pain beginning that morning. Two months ago, he was diagnosed with inferior wall myocardial infarction and was treated with stent implantation of the right coronary artery. He has a history of hypertension and hypercholesterolemia. His medications include aspirin, clopidogrel, atorvastatin, and enalapril. His temperature is 38.5 °C (101.3 °F), pulse is 92/min, respirations are 22/min, and blood pressure is 130/80 mm Hg. Cardiac examination shows a high-pitched scratching sound best heard while sitting upright and during expiration. The remainder of the examination shows no abnormalities. An ECG shows diffuse ST elevations. Serum studies show a troponin I of 0.005 ng/ml (N < 0.01). What is the most likely cause of this patient’s symptoms?

   Example Rationale 2:
   This patient had a recent myocardial infarction with new development of diffuse ST elevations, chest pain, and a high pitched scratching murmur which are found in Dressler's syndrome. This patient likely has Dressler's syndrome.

Question:

{Insert MedQA case here}”

4. Analytic reasoning chain-of-thought: “Use analytic reasoning to deduce the physiologic or biochemical pathophysiology of the patient and step by step identify the correct response.

   Example Question 1:
   Shortly after undergoing a bipolar prosthesis for a displaced femoral neck fracture of the left hip acquired after a fall the day before, an woman in her eighties woman suddenly develops dyspnea. The surgery under general anesthesia with sevoflurane was uneventful, lasting 98 min, during which the patient maintained oxygen saturation readings of 100% on 8 l of oxygen. She has a history of hypertension, osteoporosis, and osteoarthritis of her right knee. Her medications include ramipril, naproxen, ranitidine, and a multivitamin. She appears cyanotic, drowsy, and is oriented only to person. Her temperature is 38.6 °C (101.5 °F), pulse is 135/min, respirations are 36/min, and blood pressure is 155/95 mm Hg. Pulse oximetry on room air shows an oxygen saturation of 81%. There are several scattered petechiae on the anterior chest wall. Laboratory studies show a hemoglobin concentration of 10.5 g/dl, a leukocyte count of 9000/mm3, a platelet count of 145,000/mm3, and a creatine kinase of 190 U/l. An ECG shows sinus tachycardia. What is the most likely diagnosis?

   Example Rationale 1:
   The patient recently had large bone surgery making fat emboli a potential cause because the bone marrow was manipulated. Petechiae can form in response to capillary inflammation caused by fat emboli. Fat micro globules cause CNS microcirculation occlusion causing confusion and altered mental status. Fat obstruction in the pulmonary arteries can cause tachycardia and shortness of breath as seen in this patient. This patient most likely has a fat embolism.

   Example Question 2:
   A man in his fifties man comes to the emergency department because of a dry cough and severe chest pain beginning that morning. Two months ago, he was diagnosed with inferior wall myocardial infarction and was treated with stent implantation of the right coronary artery. He has a history of hypertension and hypercholesterolemia. His medications include aspirin, clopidogrel, atorvastatin, and enalapril. His temperature is 38.5 °C (101.3 °F), pulse is 92/min, respirations are 22/min, and blood pressure is 130/80 mm Hg. Cardiac examination shows a high-pitched scratching sound best heard while sitting upright and during expiration. The remainder of the examination shows no abnormalities. An ECG shows diffuse ST elevations. Serum studies show a troponin I of 0.005 ng/ml (N < 0.01). What is the most likely cause of this patient’s symptoms?

   Example Rationale 2:
   This patient had a recent myocardial infarction which can cause myocardial inflammation that causes pericarditis and Dressler Syndrome. The diffuse ST elevations and high-pitched scratching murmur can be signs of pericardial inflammation as the inflamed pericardium rubs against the pleura as seen with Dressler Syndrome. This patient likely has Dressler Syndrome

Question:

{Insert MedQA case here}”

5. Bayesian reasoning chain-of-thought: “Use step-by-step Bayesian Inference to create a prior probability that is updated with new information in the history to produce a posterior probability and determine the final diagnosis.

   Example Question 1:
   Shortly after undergoing a bipolar prosthesis for a displaced femoral neck fracture of the left hip acquired after a fall the day before, an woman in her eighties woman suddenly develops dyspnea. The surgery under general anesthesia with sevoflurane was uneventful, lasting 98 min, during which the patient-maintained oxygen saturation readings of 100% on 8 l of oxygen. She has a history of hypertension, osteoporosis, and osteoarthritis of her right knee. Her medications include ramipril, naproxen, ranitidine, and a multivitamin. She appears cyanotic, drowsy, and is oriented only to person. Her temperature is 38.6 °C (101.5 °F), pulse is 135/min, respirations are 36/min, and blood pressure is 155/95 mm Hg. Pulse oximetry on room air shows an oxygen saturation of 81%. There are several scattered petechiae on the anterior chest wall. Laboratory studies show a hemoglobin concentration of 10.5 g/dl, a leukocyte count of 9000/mm3, a platelet count of 145,000/mm3, and a creatine kinase of 190 U/l. An ECG shows sinus tachycardia. What is the most likely diagnosis?

   Example Rationale 1:
   The prior probability of fat embolism is 0.05% however the patient has petechiae on exam which is seen with fat emboli, which increases the posterior probability of fat embolism to 5%. Altered mental status increases the probability further to 10%. Recent orthopedic surgery increases the probability of fat emboli syndrome to 60%. This patient most likely has a fat embolism.

   Example Question 2:
   A man in his fifties man comes to the emergency department because of a dry cough and severe chest pain beginning that morning. Two months ago, he was diagnosed with inferior wall myocardial infarction and was treated with stent implantation of the right coronary artery. He has a history of hypertension and hypercholesterolemia. His medications include aspirin, clopidogrel, atorvastatin, and enalapril. His temperature is 38.5 °C (101.3 °F), pulse is 92/min, respirations are 22/min, and blood pressure is 130/80 mm Hg. Cardiac examination shows a high-pitched scratching sound best heard while sitting upright and during expiration. The remainder of the examination shows no abnormalities. An ECG shows diffuse ST elevations. Serum studies show a troponin I of 0.005 ng/ml (N < 0.01). What is the most likely cause of this patient’s symptoms?

   Example Rationale 2:
   The prior probability of Dressler Syndrome is 0.01%. The patient has diffuse ST elevations, increasing the probability of Dressler Syndrome to 5%. The patient has a scratching murmur which increases the probability to 10%. In the setting of a recent MI the posterior probability of myocardial infarction is 55%. This patient likely has Dressler Syndrome.

Question:

{Insert MedQA case here}”

**Full prompts used for Undiagnosed Diseases Network dataset**: All prompts are adapted from Savage, Thomas, et al. “Diagnostic reasoning prompts reveal the potential for large language model interpretability in medicine.” *NPJ Digital Medicine* 7.1 (2024): 20.

1. Traditional chain-of-thought: “Provide a step-by-step deduction that identifies the correct response.

   Example Case: “One-liner”: A teenage male with short stature, developmental delay, dysmorphic facial features, cryptorchidism, and pulmonary valve stenosis.

Category of Primary Condition: Neurology

Narrative Summary: At birth, noted to have generalized hypotonia, poor suck, and distinct facial features. Required NG feeding for the first two weeks of life. Developmental delays were evident from infancy. Rolled at 8 months, sat independently at 15 months, walked at 3 years. Now speaks only a few single words. Formal developmental assessment confirmed global developmental delay. Behavioral profile includes repetitive hand movements, mild incoordination, and attention difficulties.

Medical history notable for:

Pulmonary valve stenosis (stable)

Bilateral cryptorchidism (surgically repaired)

Recurrent otitis media (tubes x2)

Severe constipation (daily laxatives)

Percentiles:

Height: <3rd percentile

Weight: 5th percentile

Head circumference: 10th percentile

Facial Features and Measurements:

Hypertelorism (intercanthal distance: 3.6 cm, 95th percentile)

Down-slanting palpebral fissures

Ptosis

Low-set, posteriorly rotated ears (Right: 4.2 cm, Left: 4.3 cm; both <3rd percentile)

Broad/webbed neck

Shield chest with widely spaced nipples

High anterior hairline

Family History:

Father: Short stature and had a heart murmur in childhood, never fully evaluated

Paternal grandmother: Described as having a similar facial appearance

Mother: Healthy

Prior Genetic Testing:

Chromosomal Microarray: Normal

Fragile X testing: Negative

Whole Exome Sequencing (trio): VUS in PTPN11, currently under review

Karyotype: 46,XY

Known Prior Evaluations:

Brain MRI (2023): Normal

Echocardiogram: Mild-to-moderate pulmonary valve stenosis, stable

Audiology: Mild conductive hearing loss (likely secondary to otitis media)

Example Rationale: This patient is a A teenage male with short stature and global developmental delay. He had hypotonia and feeding difficulties in infancy, suggesting early neurologic involvement. His facial features, including ptosis, hypertelorism, and low-set ears, are characteristic of a syndromic condition. Additional findings such as a webbed neck, shield chest, and cryptorchidism further support this. He also has pulmonary valve stenosis, a cardiac defect commonly associated with Noonan syndrome. A VUS in PTPN11, a gene frequently implicated in Noonan syndrome, was identified. Taken together, these features point to Noonan syndrome as the most likely diagnosis.

Patient Case:

{Insert rare disease patient case here}”

2. Differential diagnosis chain-of-thought: “Use step by step deduction to create a differential diagnosis and then use step by step deduction to determine the correct response.

Example Case: “One-liner”: A teenage male with short stature, developmental delay, dysmorphic facial features, cryptorchidism, and pulmonary valve stenosis.

Category of Primary Condition: Neurology

Narrative Summary: At birth, noted to have generalized hypotonia, poor suck, and distinct facial features. Required NG feeding for the first two weeks of life. Developmental delays were evident from infancy. Rolled at 8 months, sat independently at 15 months, walked at 3 years. Now speaks only a few single words. Formal developmental assessment confirmed global developmental delay. Behavioral profile includes repetitive hand movements, mild incoordination, and attention difficulties.

Medical history notable for:

Pulmonary valve stenosis (stable)

Bilateral cryptorchidism (surgically repaired)

Recurrent otitis media (tubes x2)

Severe constipation (daily laxatives)

Percentiles:

Height: <3rd percentile

Weight: 5th percentile

Head circumference: 10th percentile

Facial Features and Measurements:

Hypertelorism (intercanthal distance: 3.6 cm, 95th percentile)

Down-slanting palpebral fissures

Ptosis

Low-set, posteriorly rotated ears (Right: 4.2 cm, Left: 4.3 cm; both <3rd percentile)

Broad/webbed neck

Shield chest with widely spaced nipples

High anterior hairline

Family History:

Father: Short stature and had a heart murmur in childhood, never fully evaluated

Paternal grandmother: Described as having a similar facial appearance

Mother: Healthy

Prior Genetic Testing:

Chromosomal Microarray: Normal

Fragile X testing: Negative

Whole Exome Sequencing (trio): VUS in PTPN11, currently under review

Karyotype: 46,XY

Known Prior Evaluations:

Brain MRI (2023): Normal

Echocardiogram: Mild-to-moderate pulmonary valve stenosis, stable

Audiology: Mild conductive hearing loss (likely secondary to otitis media)

Example Rationale: This patient has short stature, developmental delay, and congenital heart disease. The differential includes Noonan syndrome, Costello syndrome, Kabuki syndrome, and 22q11.2 deletion. The absence of coarse facies or ectodermal abnormalities makes CFC and Costello less likely. The patient lacks classic Kabuki or 22q11.2 features such as arched eyebrows or cleft palate. The presence of characteristic facial features, pulmonary valve stenosis, cryptorchidism, and a PTPN11 variant makes Noonan syndrome the most likely diagnosis.

Patient Case:

{Insert rare disease patient case here}”

3. Intuitive reasoning chain-of-thought: “Use symptoms, signs, and laboratory disease associations to step by step deduce the correct response.

Example Case: “One-liner”: A teenage male with short stature, developmental delay, dysmorphic facial features, cryptorchidism, and pulmonary valve stenosis.

Category of Primary Condition: Neurology

Narrative Summary: At birth, noted to have generalized hypotonia, poor suck, and distinct facial features. Required NG feeding for the first two weeks of life. Developmental delays were evident from infancy. Rolled at 8 months, sat independently at 15 months, walked at 3 years. Now speaks only a few single words. Formal developmental assessment confirmed global developmental delay. Behavioral profile includes repetitive hand movements, mild incoordination, and attention difficulties.

Medical history notable for:

Pulmonary valve stenosis (stable)

Bilateral cryptorchidism (surgically repaired)

Recurrent otitis media (tubes x2)

Severe constipation (daily laxatives)

Percentiles:

Height: <3rd percentile

Weight: 5th percentile

Head circumference: 10th percentile

Facial Features and Measurements:

Hypertelorism (intercanthal distance: 3.6 cm, 95th percentile)

Down-slanting palpebral fissures

Ptosis

Low-set, posteriorly rotated ears (Right: 4.2 cm, Left: 4.3 cm; both <3rd percentile)

Broad/webbed neck

Shield chest with widely spaced nipples

High anterior hairline

Family History:

Father: Short stature and had a heart murmur in childhood, never fully evaluated

Paternal grandmother: Described as having a similar facial appearance

Mother: Healthy

Prior Genetic Testing:

Chromosomal Microarray: Normal

Fragile X testing: Negative

Whole Exome Sequencing (trio): VUS in PTPN11, currently under review

Karyotype: 46,XY

Known Prior Evaluations:

Brain MRI (2023): Normal

Echocardiogram: Mild-to-moderate pulmonary valve stenosis, stable

Audiology: Mild conductive hearing loss (likely secondary to otitis media)

Example Rationale: This child has short stature, developmental delay, and characteristic facial features including ptosis, hypertelorism, and low-set ears. He also has pulmonary valve stenosis, cryptorchidism, and a webbed neck, which are findings commonly seen in Noonan syndrome. The combination of dysmorphism, congenital heart disease, and growth delay is highly suggestive. A variant in PTPN11, a gene associated with Noonan, further supports the diagnosis. This patient most likely has **Noonan syndrome**.

Patient Case:

{Insert rare disease patient case here}”

4. Analytic reasoning chain-of-thought: “Use analytic reasoning to deduce the physiologic or biochemical pathophysiology of the patient and step by step identify the correct response.

Example Case: “One-liner”: A teenage male with short stature, developmental delay, dysmorphic facial features, cryptorchidism, and pulmonary valve stenosis.

Category of Primary Condition: Neurology

Narrative Summary: At birth, noted to have generalized hypotonia, poor suck, and distinct facial features. Required NG feeding for the first two weeks of life. Developmental delays were evident from infancy. Rolled at 8 months, sat independently at 15 months, walked at 3 years. Now speaks only a few single words. Formal developmental assessment confirmed global developmental delay. Behavioral profile includes repetitive hand movements, mild incoordination, and attention difficulties.

Medical history notable for:

Pulmonary valve stenosis (stable)

Bilateral cryptorchidism (surgically repaired)

Recurrent otitis media (tubes x2)

Severe constipation (daily laxatives)

Percentiles:

Height: <3rd percentile

Weight: 5th percentile

Head circumference: 10th percentile

Facial Features and Measurements:

Hypertelorism (intercanthal distance: 3.6 cm, 95th percentile)

Down-slanting palpebral fissures

Ptosis

Low-set, posteriorly rotated ears (Right: 4.2 cm, Left: 4.3 cm; both <3rd percentile)

Broad/webbed neck

Shield chest with widely spaced nipples

High anterior hairline

Family History:

Father: Short stature and had a heart murmur in childhood, never fully evaluated

Paternal grandmother: Described as having a similar facial appearance

Mother: Healthy

Prior Genetic Testing:

Chromosomal Microarray: Normal

Fragile X testing: Negative

Whole Exome Sequencing (trio): VUS in PTPN11, currently under review

Karyotype: 46,XY

Known Prior Evaluations:

Brain MRI (2023): Normal

Echocardiogram: Mild-to-moderate pulmonary valve stenosis, stable

Audiology: Mild conductive hearing loss (likely secondary to otitis media)

Example Rationale: This patient presents with features affecting multiple organ systems: growth, neurologic development, cardiac structure, and genital development, suggesting a syndromic etiology. Pulmonary valve stenosis results from dysregulated cardiac morphogenesis during fetal development. Cryptorchidism and short stature point to disrupted Ras/MAPK signaling, which plays a key role in growth hormone response and gonadal development. The facial dysmorphisms, including hypertelorism, ptosis, and low-set ears, arise from aberrant craniofacial patterning, also mediated by the Ras/MAPK pathway. A variant in PTPN11, a key regulator in this pathway, further implicates dysregulation of intracellular signaling during development. Taken together, the clinical features and underlying pathophysiology are most consistent with **Noonan syndrome.**

Patient Case:

{Insert rare disease patient case here}”

5. Bayesian reasoning chain-of-thought: “Use step-by-step Bayesian Inference to create a prior probability that is updated with new information in the history to produce a posterior probability and determine the final diagnosis.

Example Case: “One-liner”: A teenage male with short stature, developmental delay, dysmorphic facial features, cryptorchidism, and pulmonary valve stenosis.

Category of Primary Condition: Neurology

Narrative Summary: At birth, noted to have generalized hypotonia, poor suck, and distinct facial features. Required NG feeding for the first two weeks of life. Developmental delays were evident from infancy. Rolled at 8 months, sat independently at 15 months, walked at 3 years. Now speaks only a few single words. Formal developmental assessment confirmed global developmental delay. Behavioral profile includes repetitive hand movements, mild incoordination, and attention difficulties.

Medical history notable for:

Pulmonary valve stenosis (stable)

Bilateral cryptorchidism (surgically repaired)

Recurrent otitis media (tubes x2)

Severe constipation (daily laxatives)

Percentiles:

Height: <3rd percentile

Weight: 5th percentile

Head circumference: 10th percentile

Facial Features and Measurements:

Hypertelorism (intercanthal distance: 3.6 cm, 95th percentile)

Down-slanting palpebral fissures

Ptosis

Low-set, posteriorly rotated ears (Right: 4.2 cm, Left: 4.3 cm; both <3rd percentile)

Broad/webbed neck

Shield chest with widely spaced nipples

High anterior hairline

Family History:

Father: Short stature and had a heart murmur in childhood, never fully evaluated

Paternal grandmother: Described as having a similar facial appearance

Mother: Healthy

Prior Genetic Testing:

Chromosomal Microarray: Normal

Fragile X testing: Negative

Whole Exome Sequencing (trio): VUS in PTPN11, currently under review

Karyotype: 46,XY

Known Prior Evaluations:

Brain MRI (2023): Normal

Echocardiogram: Mild-to-moderate pulmonary valve stenosis, stable

Audiology: Mild conductive hearing loss (likely secondary to otitis media)

Example Rationale: The prior probability of Noonan syndrome in the general population is low, estimated at approximately 1 in 1,000 to 1 in 2,500 live births. The presence of **short stature** and **developmental delay** increases the likelihood to around 1%. The finding of **pulmonary valve stenosis**, a cardiac lesion strongly associated with Noonan syndrome, increases the probability to 10%. Additional features including **cryptorchidism, ptosis**, **hypertelorism**, **low-set ears**, and **webbed neck** raise the probability further to 40%. A **variant of uncertain significance in** PTPN11, the most commonly mutated gene in Noonan syndrome, increases the probability to 70%. A family history of short stature and cardiac murmur in the father suggests possible inherited disease, further increasing the probability. This patient most likely has **Noonan syndrome.**

Patient Case:

{Insert rare disease patient case here}”

**Supplementary Table S1.** Summary of Repeatability Score results across models, datasets, and prompts in the empirical evaluation study. *IQR = interquartile range.*

| Model | Dataset | Prompt | Semantic Repeatability Median (IQR) | Internal Repeatability Median (IQR) |
| --- | --- | --- | --- | --- |
| ChatGPT-4 | USMLE | Traditional CoT | 0.955 (0.950-0.960) | 0.897 (0.890-0.903) |
| ChatGPT-4 | USMLE | Differential CoT | 0.945 (0.936-0.953) | 0.895 (0.888-0.901) |
| ChatGPT-4 | USMLE | Intuitive CoT | 0.958 (0.950-0.965) | 0.890 (0.883-0.897) |
| ChatGPT-4 | USMLE | Analytic CoT | 0.947 (0.939-0.954) | 0.889 (0.881-0.895) |
| ChatGPT-4 | USMLE | Bayesian CoT | 0.971 (0.967-0.975) | 0.883 (0.879-0.887) |
| ChatGPT-4 | UDN | Traditional CoT | 0.948 (0.939-0.954) | 0.886 (0.879-0.890) |
| ChatGPT-4 | UDN | Differential CoT | 0.938 (0.930-0.944) | 0.885 (0.879-0.891) |
| ChatGPT-4 | UDN | Intuitive CoT | 0.948 (0.942-0.955) | 0.880 (0.876-0.885) |
| ChatGPT-4 | UDN | Analytic CoT | 0.948 (0.941-0.954) | 0.879 (0.874-0.882) |
| ChatGPT-4 | UDN | Bayesian CoT | 0.967 (0.963-0.972) | 0.878 (0.875-0.882) |
| ChatGPT-4o-mini | USMLE | Traditional CoT | 0.955 (0.949-0.960) | 0.848 (0.838-0.858) |
| ChatGPT-4o-mini | USMLE | Differential CoT | 0.958 (0.953-0.965) | 0.897 (0.889-0.903) |
| ChatGPT-4o-mini | USMLE | Intuitive CoT | 0.958 (0.950-0.963) | 0.902 (0.896-0.906) |
| ChatGPT-4o-mini | USMLE | Analytic CoT | 0.966 (0.961-0.973) | 0.886 (0.878-0.893) |
| ChatGPT-4o-mini | USMLE | Bayesian CoT | 0.972 (0.970-0.975) | 0.826 (0.822-0.830) |
| ChatGPT-4o-mini | UDN | Traditional CoT | 0.947 (0.940-0.952) | 0.885 (0.881-0.891) |
| ChatGPT-4o-mini | UDN | Differential CoT | 0.941 (0.935-0.948) | 0.888 (0.884-0.892) |
| ChatGPT-4o-mini | UDN | Intuitive CoT | 0.948 (0.941-0.955) | 0.884 (0.878-0.892) |
| ChatGPT-4o-mini | UDN | Analytic CoT | 0.948 (0.943-0.952) | 0.884 (0.878-0.892) |
| ChatGPT-4o-mini | UDN | Bayesian CoT | 0.948 (0.944-0.954) | 0.875 (0.871-0.882) |
| Llama 3.2 1B | USMLE | Traditional CoT | 0.911 (0.904-0.918) | 0.925 (0.921-0.929) |
| Llama 3.2 1B | USMLE | Differential CoT | 0.915 (0.905-0.923) | 0.927 (0.922-0.931) |
| Llama 3.2 1B | USMLE | Intuitive CoT | 0.902 (0.895-0.909) | 0.918 (0.913-0.923) |
| Llama 3.2 1B | USMLE | Analytic CoT | 0.905 (0.897-0.912) | 0.912 (0.907-0.917) |
| Llama 3.2 1B | USMLE | Bayesian CoT | 0.915 (0.902-0.929) | 0.933 (0.926-0.939) |
| Llama 3.2 1B | UDN | Traditional CoT | 0.920 (0.912-0.929) | 0.921 (0.915-0.926) |
| Llama 3.2 1B | UDN | Differential CoT | 0.917 (0.911-0.926) | 0.922 (0.918-0.926) |
| Llama 3.2 1B | UDN | Intuitive CoT | 0.919 (0.911-0.925) | 0.918 (0.913-0.923) |
| Llama 3.2 1B | UDN | Analytic CoT | 0.920 (0.911-0.927) | 0.917 (0.911-0.921) |
| Llama 3.2 1B | UDN | Bayesian CoT | 0.911 (0.904-0.919) | 0.922 (0.918-0.927) |

**Supplementary Table S2.** Summary of Reproducibility Score results across models and datasets in the empirical evaluation study. *IQR = interquartile range.*

| Model | Dataset | Semantic Reproducibility Median (IQR) | Internal Reproducibility Median (IQR) |
| --- | --- | --- | --- |
| ChatGPT-4 | USMLE | 0.958 (0.954-0.963) | 0.992 (0.989-0.994) |
| ChatGPT-4o-mini |  | 0.960 (0.950-0.970) | 0.997 (0.993-0.998) |
| Llama 3.2 1B |  | 0.977 (0.970-0.986) | 0.990 (0.987-0.992) |
| ChatGPT-4 | UDN | 0.960 (0.957-0.963) | 0.994 (0.992-0.995) |
| ChatGPT-4o-mini |  | 0.984 (0.979-0.989) | 0.994 (0.993-0.995) |
| Llama 3.2 1B |  | 0.992 (0.989-0.994) | 0.995 (0.994-0.996) |
